# Development and Implementation of a Simple and Rapid Extraction-Free Saliva SARS-CoV-2 RT-LAMP Workflow for Workplace Surveillance

**DOI:** 10.1101/2022.03.11.22272282

**Authors:** Zhiru Li, Jacqueline L. Bruce, Barry Cohen, Caileigh V. Cunningham, William E. Jack, Katell Kunin, Bradley W. Langhorst, Jacob Miller, Reynes A. Moncion, Catherine B. Poole, Prem K. Premsrirut, Guoping Ren, Richard J. Roberts, Nathan A. Tanner, Yinhua Zhang, Clotilde K. S. Carlow

## Abstract

Effective management of the COVID-19 pandemic requires widespread and frequent testing of the population for SARS-CoV-2 infection. Saliva has emerged as an attractive alternative to nasopharyngeal samples for surveillance testing as it does not require specialized personnel or materials for its collection and can be easily provided by the patient. We have developed a simple, fast, and sensitive saliva-based testing workflow that requires minimal sample treatment and equipment. After sample inactivation, RNA is quickly released and stabilized in an optimized buffer, followed by reverse transcription loop-mediated isothermal amplification (RT-LAMP) and detection of positive samples using a colorimetric and/or fluorescent readout. The workflow was optimized using 1,670 negative samples collected from 172 different individuals over the course of 6 months. Each sample was spiked with 50 copies/μL of inactivated SARS-CoV-2 virus to monitor the efficiency of viral detection. Using pre-defined clinical samples, the test was determined to be 100% specific and 97% sensitive, with a limit of detection comparable to commercially available RT-qPCR-based diagnostics. The method was successfully implemented in a CLIA laboratory setting for workplace surveillance and reporting. From April 2021-February 2022, more than 30,000 self-collected samples from 755 individuals were tested and 85 employees tested positive mainly during December and January, consistent with high infections rates in Massachusetts and nationwide. The rapid identification and isolation of infected individuals with trace viral loads before symptom onset minimized viral spread in the workplace.

## Introduction

Early detection of infection followed by isolation of contagious individuals is key to preventing the spread of SARS-CoV-2 infection in the population. The first diagnostic tests developed involved the use of nasopharyngeal (NP) swabs to collect nasopharyngeal and oropharyngeal specimens from deep in patients’ noses and throats. This method poses considerable discomfort to the patient and requires trained healthcare professionals for sample collection. To detect virus, the NP swabs are commonly processed by purifying viral RNA upstream of reverse-transcription-quantitative polymerase chain reaction (RT-qPCR), a laborious molecular diagnostic method requiring expensive equipment and highly skilled operators. Shortages of the specialized, medical-grade NP swabs in the early days of the pandemic, as well as limited numbers of trained medical and laboratory personnel, prompted a search for alternative, simpler testing approaches that can be broadly implemented.

The use of saliva as a less invasive sampling specimen is advantageous since it can be easily self-collected into simple vessels without the need for a healthcare worker, thereby reducing a considerable bottleneck and cost. During the early and acute phases of SARS-CoV-2 infection, a relatively high viral load can be detected in saliva [1-5] and comparative studies show strong agreement in the results obtained from paired nasopharyngeal and saliva samples from the same individual [1, 6-13]. Importantly, saliva has also been shown to be an effective specimen for the detection of infection in asymptomatic individuals [10, 14, 15] who have a high rate of viral shedding [16] and therefore pose a significant threat in terms of viral spread.

Initial studies exploring the potential use of saliva primarily used RT-qPCR for SARS-CoV-2 detection [17-19]. More recently, reverse-transcription loop-mediated isothermal amplification (RT-LAMP) has emerged as an attractive and affordable alternative to RT-qPCR. RT-LAMP permits the rapid detection of pathogens without sophisticated equipment while retaining high levels of specificity and sensitivity [20, 21]. A polymerase with strand displacement activity enables exponential amplification of the target sequence under isothermal conditions [17, 20, 22]. Because of LAMP-based diagnostics simplicity, rapidity, and compatibility with various detection modalities, LAMP-based diagnostics have been deployed in low-resource or field settings, including the diagnosis and surveillance of neglected tropical diseases [23-27] and viral infections [28, 29]. During the COVID-19 pandemic, saliva-based RT-LAMP methods are increasingly being explored [8, 11, 14, 30, 31] and RT-LAMP has become a standard COVID test method alongside RT-qPCR. Compared with RT-qPCR, the tolerance of the reaction chemistry used in LAMP to the inhibitors present in clinical samples [21, 32] can obviate the need for a nucleic acid extraction/purification step, reducing both the time and cost to process samples. In the case of saliva, a more biologically complex sample [33] than nasal fluid, additional care should be taken when a minimal or extraction-free method is being considered. Saliva pH, color, viscosity, and RNAse activities can vary widely and potentially impact the ability to detect viral RNA.

In the present study, we report an extraction-free, saliva-based RT-LAMP workflow for SARS-CoV-2 detection with the option of a simple colorimetric endpoint and/or a semi-quantitative fluorescence readout. We demonstrate the robustness of the method using a large cohort of contrived human samples and successful implementation for frequent surveillance testing in the workplace. This has enabled us to identify and isolate infected individuals with trace viral loads before symptom onset and limit viral spread.

## Materials and Methods

### Ethical approval

Review and approval were obtained from the WCG/New England Institutional Review Board (IRB), Study Number 1298746. All relevant ethical guidelines and regulations were followed and study participants, or their legal guardians, provided written informed consent. Collection, processing, and reporting of COVID-19 testing results were performed in compliance with the CLIA (Clinical Laboratory Improvement Amendments) registration granted to New England Biolabs.

### Saliva samples

A total of 1,670 saliva samples were donated by 172 uninfected individuals during October 2020-March 2021 as part of the IRB study. Each specimen was anonymized prior to use. Between April 2021-February 2022, 755 individuals participated in a workplace surveillance program, providing saliva specimens 1-3 times/week. SARS-CoV-2 reference positive and negative saliva samples, previously tested following an RNA extraction and RT-qPCR procedure, were kindly provided by Mirimus Clinical Labs (Brooklyn, NY).

Prior to saliva collection, donors were requested to refrain from drinking anything but water, eating, chewing gum or tobacco, or smoking for at least 30 minutes prior to collection. Saliva was self-collected by passive drooling through a 1.0 mL unfiltered pipette tip into 1.5 mL tubes, each pre-applied with a pair of QR Codes, one on the top and one on the side, for accurate specimen identification. Unless specified, samples were stored at room temperature for less than 4 hours or overnight at 4°C prior to testing. Tubes containing saliva were heated at 65°C for 30 min in a benchtop incubator (Boekel Scientific, Model 138200) before opening to inactivate any virus present [34]. All samples were handled in a biohazard hood (SterilGARD Hood Class II Type A/B3).

### Sample preparation

An extraction-free saliva lysis buffer (SLB) was developed and prepared as a 2X solution containing 5 mM tris(2-carboxyethyl) phosphine (TCEP, Millipore Cat# 580567), 22 mM sodium hydroxide (Sigma 72068), 2 mM Ethylenediaminetetraacetic acid (EDTA, Invitrogen 15575-038) and 0.4% Pluronic F-68 (Gibco 24040-032). To release and stabilize RNA for detection, 15 μL of heat-inactivated saliva samples were mixed with 15 μL of 2X SLB buffer. Mixed samples were heated in a thermocycler (Bio-Rad T100 Thermal Cycler) at 95°C for 5 min, unless otherwise indicated, then cooled to 4°C. For each RT-LAMP reaction, 2 μL of treated saliva sample was used in a 20 μL reaction.

For comparison studies, RNA was purified from saliva using the Monarch® Total RNA Miniprep Kit (NEB, Cat# T2010) following the protocol for Saliva, Buccal Swabs, and Nasopharyngeal Swabs. RNA was eluted with 80 μL of nuclease-free water, quantified using the Qubit RNA BR Assay Kit (ThermoFisher Scientific, Cat # Q33224) and stored at -80°C.

### Control Virus and RNA

As a positive control, gamma-irradiated (BEI, Cat# NR-52287, Lot No. 70035888) or heat-inactivated (BEI, Cat# NR-52286, Lot no. 70034991) SARS-CoV-2 virions were used to spike saliva. The reported genome copies for γ-irradiated and heat-inactivated SARS-CoV-2 were 1.75×10^9^ and 3.75×10^8^ genome equivalents/mL, respectively. Viral stocks were aliquoted into small volumes and stored at -80°C until use. Diluted viral stock solutions were also prepared in a freezing solution (10% glycerol, 2.5% ethylene glycol) and stored at -80°C. To generate a dilution series, a known amount of virus was serially diluted in saliva obtained from SARS-CoV-2 negative donors. Synthetic SARS-CoV-2 RNA (Twist Bioscience, Cat # MN908947.3) was used to generate a standard curve for RT-qPCR. Total human RNA from HeLa cells (ThermoFisher, Cat# AM7852) was used to validate reagents. RNAs were aliquoted and stored at -80°C.

### RT-qPCR

RT-qPCR was performed using the Luna® SARS-CoV-2 RT-qPCR Multiplex Assay Kit (NEB Cat # E3019) following the manufacturer’s instructions. Each 20 μL reaction contained 2 μL RNA purified from saliva. N1 (HEX), N2 (FAM) and RNase P (Cy5) targets were simultaneously detected using the following cycling conditions: carryover prevention (25°C for 30 sec), cDNA synthesis (55°C for 10 min), initial denaturation (95°C for 1 min) followed by 45 cycles of alternating denaturation (95°C for 10 sec) with annealing/elongation (60°C for 30 sec) plus a plate read step.

### RT-LAMP

RT-LAMP reactions were performed using the SARS-CoV-2 Rapid Colorimetric LAMP Assay Kit (NEB Cat # E2019), targeting N and E regions of the SARS-CoV-2 genome, according to the manufacturer’s instructions. For each specimen, an endogenous actin control reaction was performed to ensure that sample lysis was achieved, and the saliva was of sufficient quality. Each sample was tested for both SARS-CoV-2 (COVID LAMP) and actin (Actin LAMP), unless otherwise stated. Reactions containing water served as no template controls (NTC) to monitor for any background signal. Inactivated SARS-CoV-2 virus was used as a positive control to ensure the proper functioning of the COVID LAMP reaction. Human RNA was used as a positive control for the actin LAMP reaction. To enable reaction dynamics to be monitored in real-time, SYTO™ 9 green fluorescent nucleic acid stain (Invitrogen Cat # S34854) was added to a final concentration of 1 μM in the colorimetric LAMP reaction. Each 20 μL reaction in a strip tube or 96-well plate was run at 65°C in a Bio-Rad CFX96 or Opus thermocycler and fluorescence was read in the SYBR/FAM channel every 15 seconds for 97 “cycles”. The total reaction time was ∼35 minutes with each “cycle” corresponding to ∼22 seconds reaction time (15 seconds combined with plate reading time). Following completion of RT-LAMP, data were processed using the Bio-Rad CFX-Maestro software using baseline subtracted curve fit and fluorescent drift correction. The time (min) to reach the fluorescence detection threshold was determined (Tt). No amplification is denoted N/A or assigned a Tt of 36 minutes for plotting purposes. To record color changes, tubes or 96-well plates were imaged using an Epson Perfection V600 Photo Scanner before and after the RT-LAMP reaction. A Tt ≤ 26 minutes with a post amplification color of yellow indicated the detection of target, whereas a Tt > 26 minutes or N/A, and a post reaction color of pink, indicated no detection.

### Extraction-free saliva RT-LAMP workflow for workplace surveillance

Employee samples were processed in a CLIA-registered facility at New England Biolabs. Saliva was collected in 1.5 mL QR coded tubes and self-registered in a purpose-built Laboratory Information Management System (LIMS) using cell phone QR code recognition or manually entered codes. Registered tubes were placed in 96-well microcentrifuge tube racks. Images of the loaded racks, with positive and negative controls added, were taken with a mounted cell phone camera by the lab operator and uploaded to LIMS to track sample location and generate a plate map file for import into the Bio-Rad CFX-Maestro software. Following heat treatment at 65°C for 30 minutes to inactivate any virus present, 15 μL of sample was mixed with an equal volume of SLB buffer in barcoded 96-well plate and heated at 95°C for 5 minutes to release and protect viral RNA for detection. An aliquot of 2 μL saliva lysate was then transferred to a 96-well barcoded plate containing the RT-LAMP reaction mix for viral detection and a second barcoded plate for endogenous actin detection. Saliva was tested within 4 hours of collection and both color and real-time fluorescent readouts were obtained for each sample.

A negative COVID test result was reported when the COVID LAMP assay was negative with endogenous actin detected. If actin was not detected, the sample was retested in triplicate and reported as inconclusive if any of the repeat testing failed to detect actin. When a positive COVID LAMP result was obtained, repeat testing was also performed in triplicate. A final determination of a positive COVID result required 2 or more positive results in repeat testing, otherwise, the sample would be scored inconclusive (1/3 positive) or negative (0/3 positive).

The pre- and post-amplification images of the colorimetric reactions as well as amplification files were uploaded and stored in the LIMS tracking system. The test result was automatically generated from the database based on the cutoff values and criteria described above. All automated data was manually reviewed before release of results to individuals or to state health agencies. Links to results were released to employees via automated email immediately if negative, or after repeat testing in triplicate for confirmation of a positive or inconclusive outcome. The processing time from sample inactivation to reporting results was ∼2 hours.

## Results

### Optimization of a sample preparation method for rapid release and stabilization of viral RNA

To achieve highly reliable detection of SARS-CoV-2 in crude saliva, several buffer formulations including both commercial and previously published protocols were evaluated (data not shown) before selection of a commonly used buffer containing TCEP, NaOH, and EDTA [35]. This buffer was subsequently modified to include the detergent Pluronic F-68 and referred to as saliva lysis buffer (SLB). The performance of SLB (with and without Pluronic F-68) was evaluated on contrived saliva samples. A pool of fresh saliva from uninfected donors (10 individuals) was spiked with 10-40 copies of virus /μL (6 replicates). Both real-time fluorescence (Tt) and colorimetric (pink/negative, yellow/positive) readouts were obtained for each sample (**Figure 1A**). The inclusion of Pluronic F-68 enabled more consistent detection of virus, particularly when less virus was present. At 40 copies/μL, all samples turned from pink to yellow in the presence of Pluronic F-68, whereas 1/6 remained pink in the absence of detergent, indicating a failure to detect the virus in this sample. At the lowest concentration tested, 10 copies/μL, 5/6 replicates scored positive using SLB, in contrast to only 3/6 when no detergent was present. A similar trend was observed using fluorescence as the output with all positive (yellow) samples reaching the detection threshold between 10-15 minutes, whereas the samples that scored negative (pink) had very delayed Tt or none (N/A). The detergent did not interfere with the quality of the color readout, nor the ability to detect endogenous actin in the same specimen (Actin LAMP). Actin was easily detected with or without Pluronic F-68.

**Figure 1.**
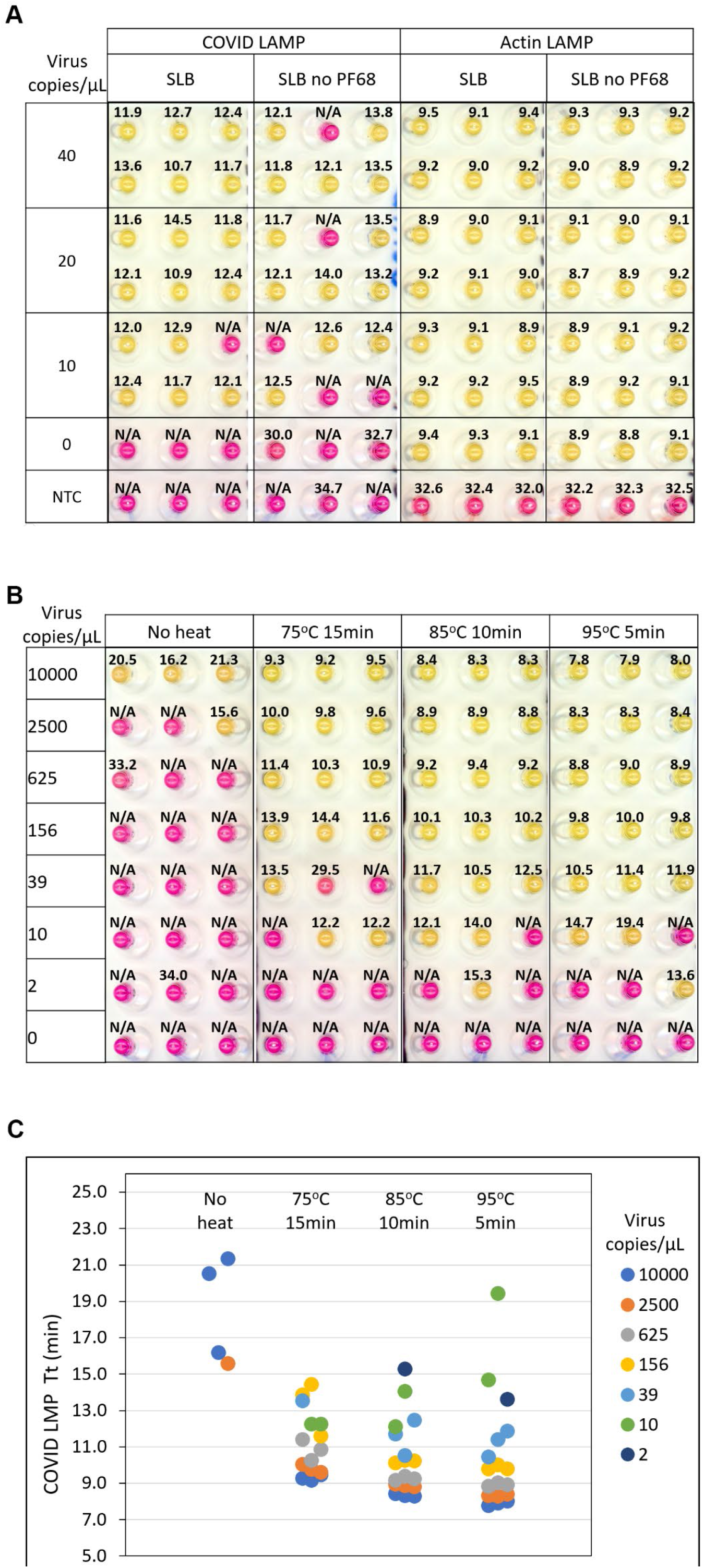
The effects of the detergent Pluronic F68 and heat treatment on viral detection in saliva. **(A)** Contrived saliva samples were processed in SLB with or without detergent followed by heat treatment (95°C for 5 min). Samples were spiked with 10-40 copies of virus /μL (6 replicates). Both real-time fluorescence (Tt value) and colorimetric (pink/negative, yellow/positive) readouts were obtained for each sample. The scanned image of the post-amplification plate (COVID LAMP and actin LAMP) is shown with overlaid Tt values. **(B)** Contrived samples spiked with 2-10,000 viral copies/μL, were processed in triplicate using either 75°C for 15 min, 85°C for 10 min, 95°C for 5 min or no heat. The scanned image of the post amplification plate (COVID LAMP) is shown with overlaid Tt values. **(C)** The plot of Tt values from (B). No amplification is denoted N/A.

To evaluate the performance of SLB at various temperatures, contrived samples spiked with 2-10,000 viral copies/μL, were processed in triplicate using either 75°C for 15 min, 85°C for 10 min or 95°C for 5 min (**Figure 1B-C)**. It was apparent that heat treatment was essential with the highest levels of sensitivity achieved after heating at 85°C for 10 min or 95°C for 5 min, both detecting 39 copies/μL in all triplicate samples. Heating samples at 95°C for 5 min resulted in the best performance and greatest reproducibility across a range of viral loads and was selected as the optimal temperature to rapidly release and stabilize viral RNA. This protocol also proved optimal for actin detection (**Supplementary Figure S1**).

### Impact of saliva input on diagnostic sensitivity

Saliva is a complex biological mixture that can vary widely in pH, color, and viscosity. These factors can impair amplification efficiency and influence the performance of the test [36]. The characteristics that affect saliva viscosity include the presence of aggregates, variations in temperature, and the time elapsed between sample collection and testing. Saliva naturally settles into supernatant and sediment phases soon after collection and storage. To determine if the virus is associated with a particular phase and if the performance of RT-LAMP is impacted by the different phases, supernatant, sediment, and evenly resuspended sample from crude positive saliva were each evaluated as input material (**Figure 2A**). Virus was detected in all phases with the fastest amplification (lowest Tt) observed in the supernatant, indicating that saliva supernatant is optimal for testing and no pre-mixing is required. Viral and actin detection was least efficient when crude sediment was used as input. For comparison, RNA was also purified from supernatant and sediment. This resulted in overall faster amplification for both COVID and actin assays than with crude lysate, likely due to the elimination of inhibitors or interfering substances present in crude saliva. These results demonstrate that saliva supernatant is suitable for viral detection, obviating the need for an additional re-suspension step, which is particularly helpful when dealing with large numbers of samples simultaneously.

**Figure 2.**
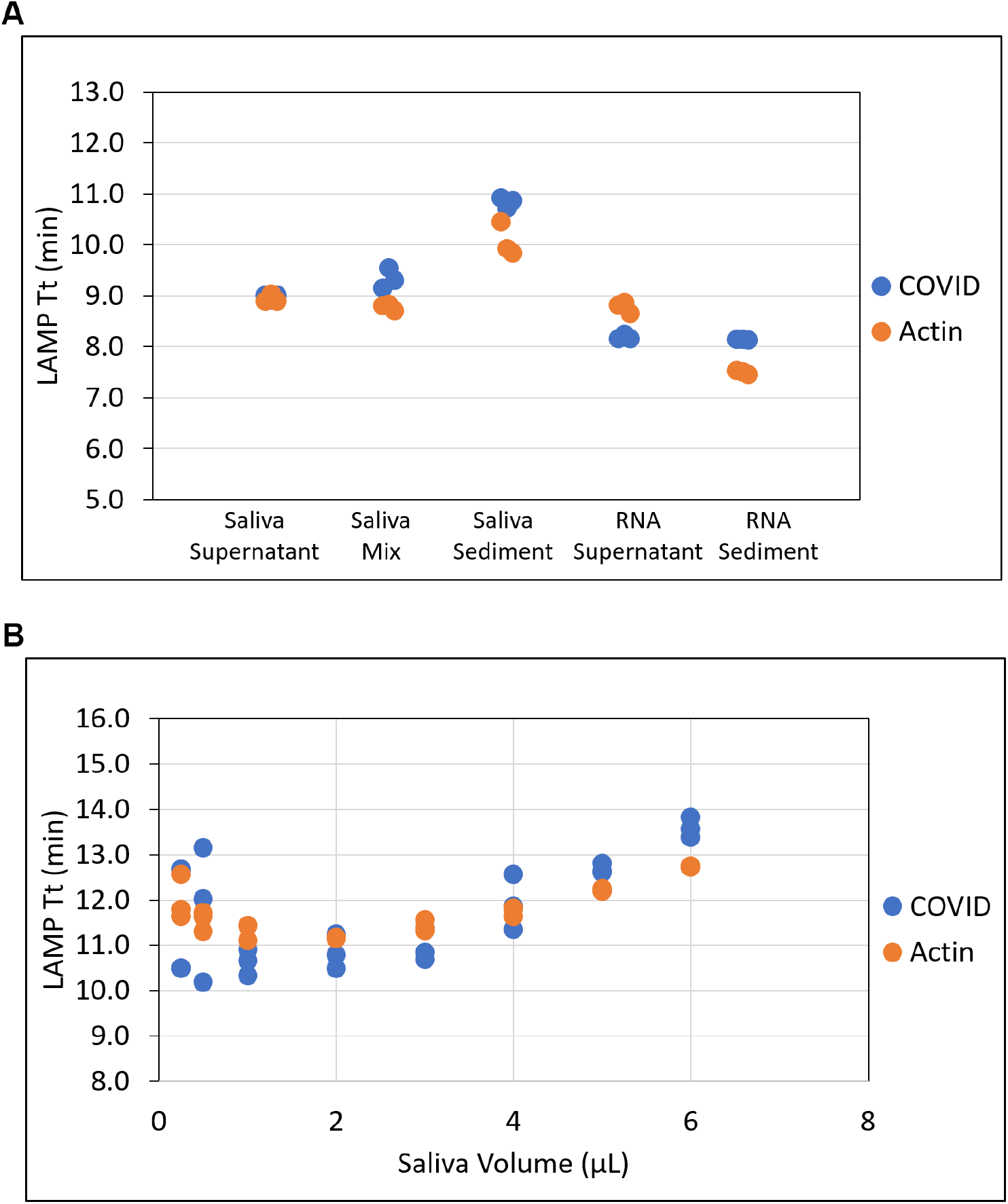
Impact of saliva input on diagnostic sensitivity. **(A)** Supernatant and sediment from positive saliva and corresponding purified RNAs were evaluated in COVID and actin LAMP reactions. **(B)** Different volumes of positive saliva ranging from 0.25 μL to 6 μL were used in COVID and actin LAMP reactions. LAMP Tt values were plotted. All samples were tested in triplicate.

To evaluate the tolerance of the LAMP assay to increasing amounts of saliva, amounts ranging from 0.25-6 μL SARS-CoV-2 positive saliva were used as input in a 20 μL reaction (**Figure 2B**). At all volumes of saliva input, virus was detected, but the best overall performance was observed with 1 μL of a saliva sample. Lower than 1 μL, more variation was observed in triplicate samples. Increasing saliva input volume did not increase sensitivity and delayed the reaction time for viral detection, with a difference as much as 4 min when using 1 μL versus 6 μL saliva. A similar trend, though not as pronounced, was also observed with respect to actin detection following volumetric adjustments in saliva input (**Figure 2B)**.

To evaluate the robustness of the extraction-free LAMP assay to individual variation and optimize the workflow, 1,670 negative saliva samples were collected from 172 different individuals over the course of 6 months. This diverse set of saliva specimens was used to generate contrived samples each spiked with 50 copies/μL. Of the 1,670 spiked samples, 1,646 (98.6%) scored positive in both colorimetric (data not shown) and fluorescent LAMP assays (**Figure 3**) using 26 minutes as a cutoff. Most (91.7%) samples scored positive for COVID within 15 minutes using the real-time fluorescent signal, whereas 24 samples failed to amplify indicating a false negative rate of 1.4% (**Figure 3A)**. In the actin control LAMP, 1,664 (99.6%) scored positive, most of them (95.6%) within 15 minutes (**Figure 3B**). When the Tt values from COVID and actin amplification reactions from all the samples were plotted, a positive correlation (Pearson correlation coefficient R^2^ value of 0.63, P-value < 0.0001) was observed (**Figure 3C**).

**Figure 3.**
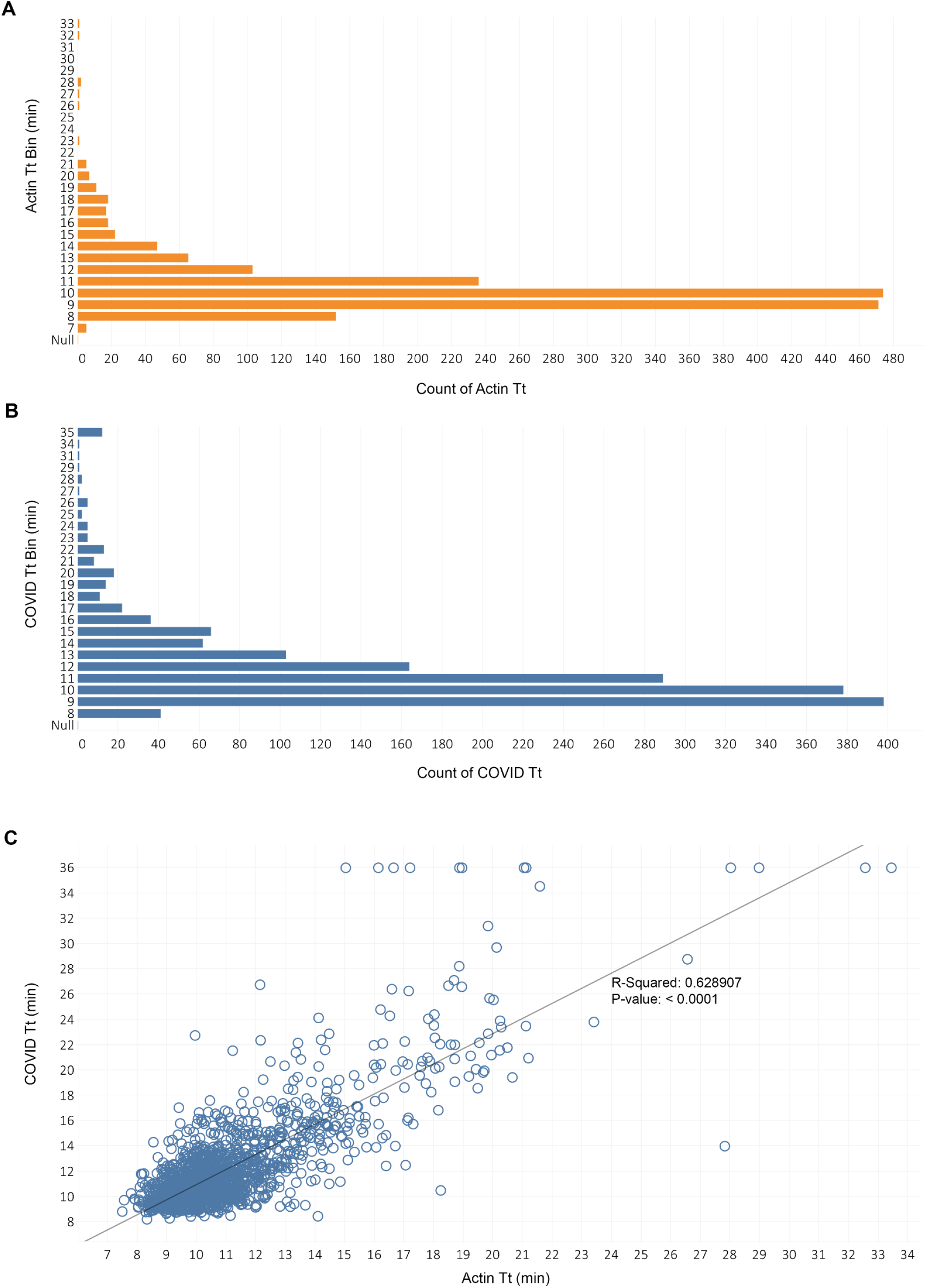
Performance of the extraction-free RT-LAMP method using saliva collected from a large cohort of individuals. A total of 1670 negative saliva samples were each spiked with 50 copies/μL of virus and evaluated in COVID and actin LAMP reactions. Histogram depicting the distribution of Tt values from COVID **(A**) and actin (**B**) LAMP reactions are shown. (**C**) Plotted COVID and actin Tt values (blue circles) and linear regression line with the value of R squared (Pearson product-moment correlation coefficient) and p-value. No amplification is assigned a Tt value of 36 for plotting purposes.

Pooling saliva is often an attractive option to minimize testing costs. To assess the suitability of the method for pooled saliva testing, one SARS-CoV-2 positive saliva specimen was combined with equal volumes of 1 to 15 randomly selected negative saliva samples from different individuals, corresponding to dilution series of 1:2 through 1:16 of the original viral titer. All 15 sample pools and the single positive specimen yielded a positive result (**Supplementary Figure S2**). In this experiment, problematic samples which contained inhibitory substances were diluted in the pool, indicating that a pooling strategy can be beneficial in cases where a particular specimen has a low pH, is colored or viscous, or contains salivary inhibitors impairing amplification efficiency.

### Clinical performance of the extraction-free saliva RT-LAMP workflow

Taking into consideration the variability observed between individual saliva specimens, two separate pools were each generated by combining equal volumes of saliva from ten different negative individuals and used to determine the limit of detection (LoD). Twenty replicate samples were prepared from each pool and spiked with 40 copies of virus /μL and then tested in triplicate. All 60 reactions from each of the 2 pools (**Figure 4**) scored positive (100% sensitivity) within 10 (pool 1) or 11 (pool 2) minutes. When spiked with fewer copies of virus, the viral RNA was also detected but at a reduced frequency. At 20 copies/μL or 10 copies/μL, sensitivities of 82% (49/60) and 63% (38/60) were obtained, respectively. Therefore, the LOD for this direct saliva RT-LAMP assay is 40 copies of virus/μL of saliva.

**Figure 4.**
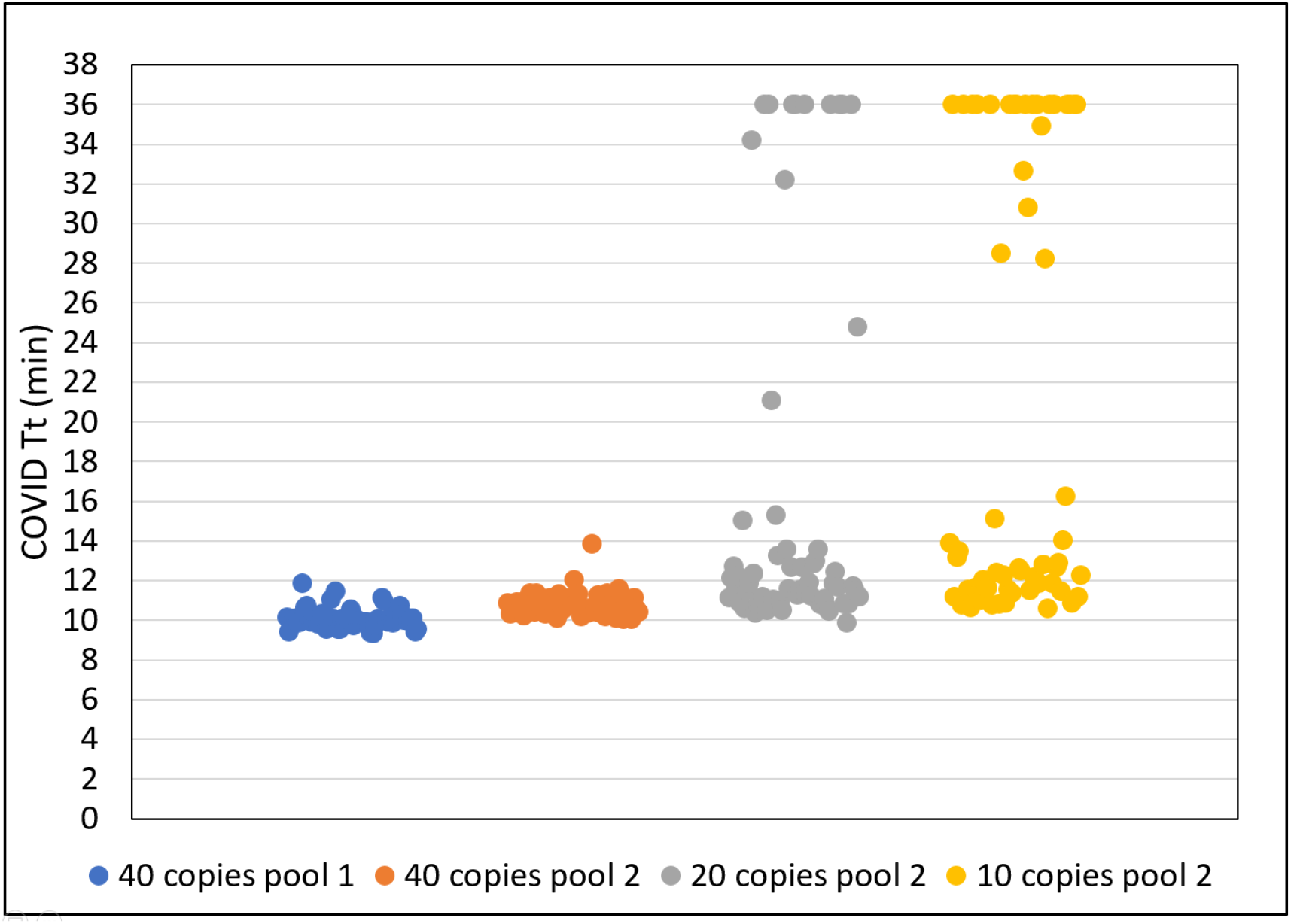
The limit of detection. Two saliva pools from different negative individuals (n=10) were spiked with 10-40 copies of virus /μL. Three LAMP reactions were performed using 20 different replicate samples. The Tt values derived from 60 reactions are plotted. No amplification is assigned a Tt value of 36 for plotting purposes.

To evaluate the diagnostic capabilities of the workflow using pre-defined clinical samples, a total of 30 positive and 30 negative saliva specimens were tested at least three times by two different operators in a blinded manner (**Supplementary Figure S3**). The status of these samples had previously been determined in an RT-qPCR test using purified RNA from each sample as input (Mirimus Inc.). The overall clinical sensitivity [(True Positives)/(True Positives + False Negatives)] was 97% and specificity [(True Negatives)/(True Negatives + False Positives)] of 100%, demonstrating the high accuracy of the workflow.

To compare the performance of RT-LAMP with RT-qPCR, 16 saliva samples containing a range of viral load were tested in both assays. For RT-LAMP, both saliva lysate and purified RNA were used as input, whereas RT-qPCR was performed using only purified RNA (**Table 1**). Cq values in RT-qPCR, ranged from 24-39, and all 11 samples with Cq values less than 35 also tested positive in RT-LAMP using saliva lysate as input. When purified RNA was used in RT-LAMP, one additional sample (#12) scored positive. This sample had a very low viral load with a Cq value greater than 35 in RT-qPCR, equivalent to ∼ 5 copies of viral RNA (**Supplementary Figure S4**). These data demonstrate the high performance of the extraction-free saliva RT-LAMP workflow, which is considerably simpler and faster to perform than RT-qPCR.

**Table 1.** Direct comparison of SARS-CoV-2 detection using RT-qPCR and extraction-free RT-LAMP.

| Samples | Purified Saliva RNA |  |  |  |  |  | Saliva Lysate |  |
| --- | --- | --- | --- | --- | --- | --- | --- | --- |
|  | RT-qPCR N1<br>(Cq) |  | RT-qPCR N2<br>(Cq) |  | COVID LAMP<br>(Tt) |  | COVID LAMP<br>(Tt) |  |
| 1 | 24.8 | 24.8 | 24.2 | 24.3 | 8.9 | 8.9 | 8.7 | 8.6 |
| 2 | 26.2 | 26.3 | 25.8 | 25.8 | 9.9 | 9.9 | 9.8 | 10.1 |
| 3 | 27.3 | 27.2 | 26.9 | 27.0 | 8.5 | 8.4 | 10.7 | 10.3 |
| 4 | 28.6 | 28.7 | 28.2 | 28.2 | 9.5 | 9.6 | 9.4 | 9.4 |
| 5 | 30.1 | 30.2 | 29.8 | 29.8 | 9.2 | 8.9 | 10.7 | 10.3 |
| 6 | 31.6 | 31.7 | 31.1 | 31.1 | 9.4 | 9.2 | 13.3 | 11.5 |
| 7 | 31.7 | 32.0 | 31.2 | 31.6 | 10.3 | 9.8 | 11.3 | 11.0 |
| 8 | 32.0 | 32.0 | 31.3 | 31.3 | 9.5 | 9.6 | 12.0 | 11.9 |
| 9 | 32.9 | 33.3 | 36.1 | 36.5 | 12.1 | 12.0 | 13.6 | 13.6 |
| 10 | 34.3 | 34.5 | 33.4 | 34.0 | 10.3 | 10.9 | 10.6 | 11.6 |
| 11 | 34.7 | 34.7 | N/A | N/A | 14.0 | 13.3 | 16.5 | 14.7 |
| 12 | 35.3 | 35.5 | 35.4 | 35.2 | 12.0 | 11.7 | 22.9 | N/A |
| 13 | 37.5 | N/A | N/A | N/A | N/A | N/A | 16.4 | N/A |
| 14 | 37.8 | 37.5 | 37.9 | 38.3 | 13.8 | 28.4 | N/A | N/A |
| 15 | 38.4 | 38.1 | N/A | N/A | N/A | 19.8 | N/A | N/A |
| 16 | 38.8 | 39.1 | N/A | N/A | N/A | N/A | N/A | N/A |

No amplification is denoted N/A.

### Successful implementation of the extraction-free saliva RT-LAMP workflow for workplace surveillance

A workflow from specimen collection to testing and result reporting was implemented in a CLIA lab setting (**Figure 5, Supplementary Figure S5)**. During the 45-week period, employees were tested once per week from April to July 2021, and from August 2021 to February 2022, the frequency was increased to twice per week. A total of 755 individuals (registered as 406 males, 341 females, and 8 gender not indicated) provided 32,906 self-collected saliva samples for testing. The highest number of positive cases was observed between 24 December 2021 and 14 January 2022 (**Figure 6A**). Of the 85 positive individuals, there were 47 males, 35 females, and 3 gender not indicated. Samples from males scored positive within 7-23 minutes (**Figure 6B**). While the difference between Tt derived from males and females is not statistically significant (Wilcoxon ranked sum, p value = 0.075), only samples from males (9) were observed with Tt ≤8 indicating a high virus titer in these specimens. In analyzing the testing history of positive individuals, we found at least 13% of positive individuals still testing positive after 10 days, however, most of these were negative by Day 14 (**Figure 6C**). No difference in the rate of viral clearance in males versus females was observed. Importantly, only 42/32,906 samples generated an inconclusive result (0.13%) after triplicate repeats, reflecting both the robustness of the test and the quality of the samples submitted for testing.

**Figure 5.**
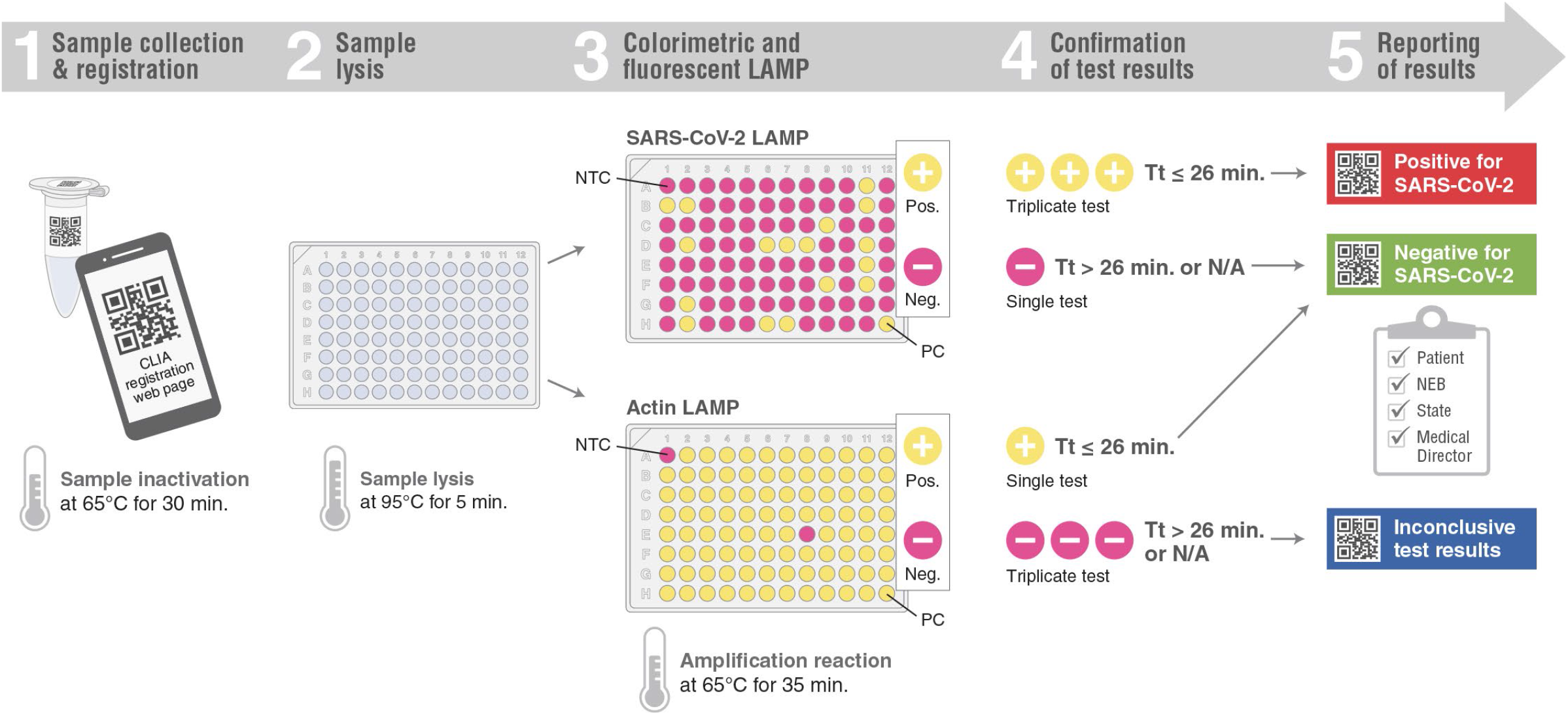
**Diagrammatic representation of the extraction-free saliva SARS-CoV-2 RT-LAMP workflow process from specimen collection to testing and result reporting.**

**Figure 6.**
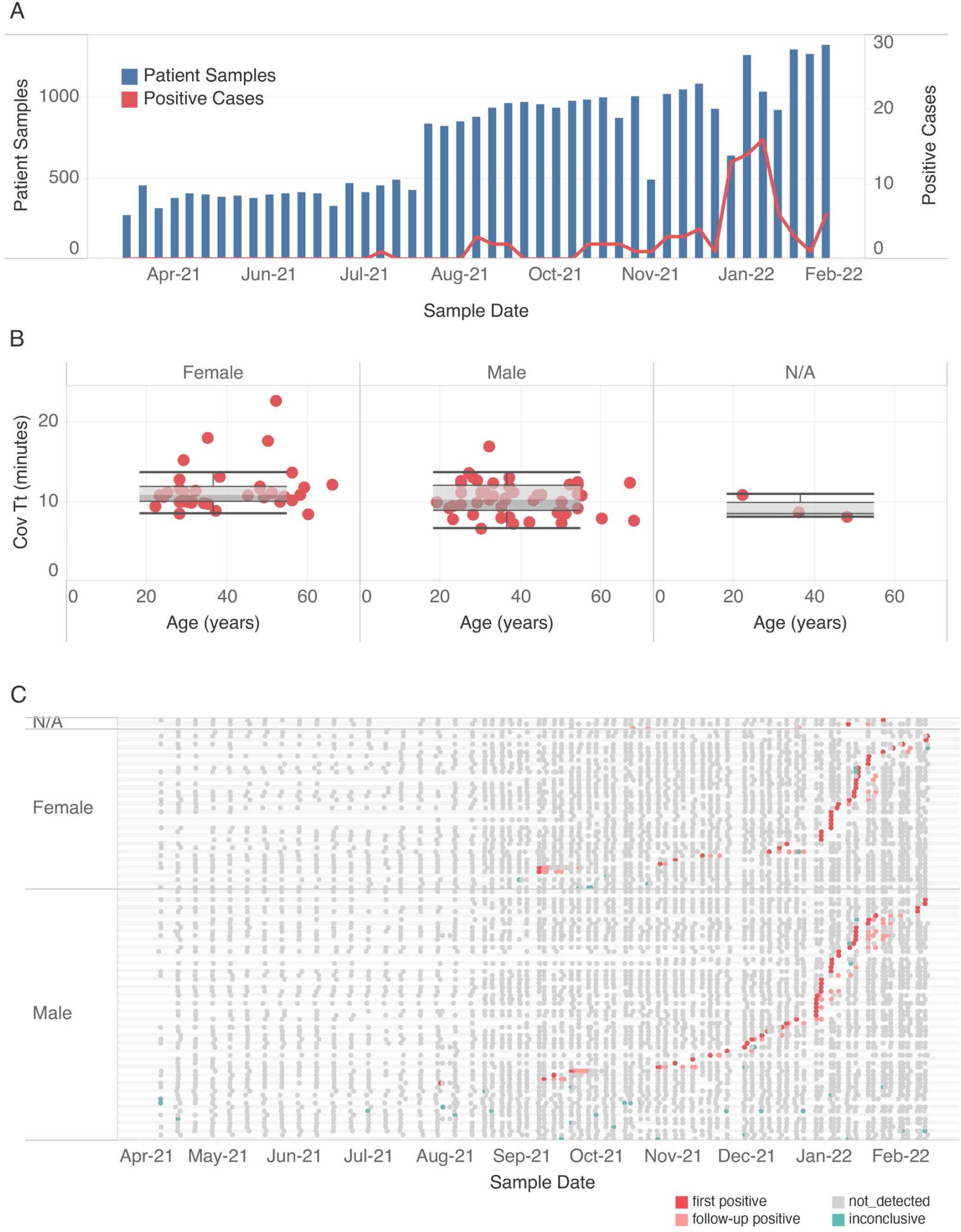
Using extraction-free saliva RT-LAMP for workplace surveillance. **(A)** The number of samples tested each week from 4/9/2021 until 2/9/2022 (blue bars) and new positive cases identified (red line). **(B)** Distribution of initial COVID Tt values obtained from male, female and individuals with gender not indicated (N/A). Average Tt values for first positive cases (Y-axis) based on gender (columns) and submitter age (X-axis), median (line), quartile boundaries (boxes), and 1.5x the interquartile range (whiskers) are displayed. **(C)** Testing history of all positive cases from April 2021 to February 2022. For each individual, negative (grey), positive (dark red, first detected; light red, subsequent detection), or inconclusive (green) results are indicated.

To compare the two readouts used, namely the fluorescence versus an endpoint color determination, a blinded study was performed using 3,654 (∼10% of total specimens processed) samples collected from 18 January 2022 to 4 February 2022. Without prior knowledge of the results (157 positives, 3,486 negatives, and 11 inconclusives), the endpoint color readout was scored by 3 different operators, taking into consideration both COVID and actin reactions (**Table 2**). A strong agreement between the two readout methods was observed with a yellow color (positive) corresponding to a fast amplification and pink/orange shaded as negative (Tt > 26 minutes or N/A). The six samples that were not detected by eye, were orange shaded and corresponded with late Tt values, representing low viral loads or at the limit of detection of the test. This demonstrates the reliability of the visual inspection method and the ability to use colorimetric RT-LAMP for simplified diagnostic workflows.

**Table 2.** Comparison of the two assay readouts: fluorescence versus color assessment.

|  |  | **Positive** | **Negative** | **Inconclusive** |
| --- | --- | --- | --- | --- |
| **Fluorescence** | **COVID Tt** | $\leq 26$ min | $> 26$ min | $> 26$ min or N/A |
| | **Actin Tt** | $\leq 26$ min | $\leq 26$ min | $> 26$ min or N/A |
|  | **Sample Number** | 157 (4.3%) | 3486 (95.4%) | 11 (0.3%) |
| **Color** | **COVID** | Yellow | Pink/Orange | Pink/Orange |
|  | **Actin** | Yellow | Yellow | Pink/Orange |
|  | **Operator 1** | 149 (4.1%) | 3497 (95.7%) | 8 (0.2%) |
|  | **Operator 2** | 152 (4.2%) | 3491 (95.5%) | 11 (0.3%) |
|  | **Operator 3** | 152 (4.2%) | 3496 (95.7%) | 6 (0.2%) |

A blinded study was performed on 3,654 samples using two readouts, namely fluorescence and endpoint color. The endpoint color readout was scored by 3 different operators. The number of samples (and percent of total) scored as positive, negative, or inconclusive are shown.

## Discussion

The goals of the present study were to develop a simple, rapid, and robust SARS-CoV-2 workflow for saliva testing and implement the method for workplace surveillance where frequent testing of asymptomatic individuals is required to prevent transmission.

While several saliva-based approaches have been demonstrated for the detection of SARS-CoV-2 using LAMP [8, 14, 30, 31, 35, 37-41], including extraction-free or direct protocols [13, 30, 35, 37, 38, 40, 42, 43], no gold standard method exists. We focused on the development of a simple and improved sample preparation protocol compatible with this complex sample and downstream LAMP reactions. We examined supernatant and sediment fractions which naturally form within minutes of collection in a crude saliva specimen and determined that the virus is present and easily detectible in both, circumventing the need for a mixing step prior to sampling. Following collection, the virus is known to be stable in saliva for extended periods of time at elevated temperatures without the addition of preservatives [18, 44, 45]. However, once RNA is released from the viral envelope, we found that it is rapidly degraded if not protected. This is likely due to the high level of endogenous RNases in human saliva [46, 47]. In the extraction-free method described in this study, 95°C heating is used to disassemble the viral particle and release RNA as well as denature and inactivate some of the RNases in the presence of a buffer containing a TCEP/EDTA mixture. TCEP is a reducing reagent which abolishes the activity of RNases via the reduction of disulfide bonds present in the enzyme, while EDTA chelates the divalent cations required for nuclease activity. The non-ionic surfactants Tween 20 and Triton X-100 have also been used to help disrupt the viral envelope and release RNA upstream of RT-LAMP [35]. However, we observed a rapid degradation of viral RNA and failure to detect virus when positive saliva samples were incubated at room temperature for 5-10 minutes in buffer containing Tween 20 and Triton X-100 (data not shown). Indeed Tween-20 and Triton X-100, used in the range of 0.1-2.0%, have been reported to increase human RNase activity [46]. We screened a panel of surfactants (data not shown) and discovered that Pluronic F-68 did not enhance the endogenous RNase activity present in saliva samples and demonstrated that the addition of 0.2% Pluronic F-68 to the sample preparation buffer increased the sensitivity of the RT-LAMP assay. Pluronic F-68 is an environmentally friendly [48], non-ionic detergent, commonly used to reduce foaming in stirred cultures and reduce cell attachment to glass. Detergents dissolve the lipid bilayer by forming a micelle, a process that depends on temperature and the critical micelle concentration (CMC). While Triton X-100 has a low CMC of ∼0.02% (0.02g/dL) at room temperature, Pluronic F-68 has a substantially higher CMC of 10g/dL at 20°C, but a much lower one at a higher temperature 0.5g/dL at 50°C [16, 49]. The temperature-dependent CMC of Pluronic F-68 may maintain the integrity of the virus at low temperature while the RNase activity is still high and subsequently facilitate the release of RNA during the heating step when the RNase activity is lower.

The constituents of saliva vary significantly both within and between individuals and are subject to collection method, hydration, and circadian rhythms [50]. Saliva has a pH normal range of 6.2-7.6 [51]. Colorimetric LAMP uses the pH-sensitive dye phenol red which turns from pink/red to yellow at pH 6.8 and lower, following the generation of hydrogen ions resulting from amplification of the target [52-54]. Low pH saliva can cause reactions to instantaneously change to yellow pre-amplification and, if not noted, will result in a false positive determination [29, 36]. In one study, 7% of saliva samples tested triggered a color change without amplification [11], and in another 15% of specimens showed a discordant color output when compared with an agarose gel electrophoresis readout [30]. In the present study we demonstrated the robustness of the lysis system when challenged with more than 1670 spiked crude saliva samples, with only 0.13% of samples invalidated due to a color change pre-amplification. For these highly acidic samples, the addition of sodium hydroxide, to a final concentration of 30mM, usually corrected the pre-amplification color to pink and enabled actin detection and validation of the corresponding COVID test (data not shown). Since low pH saliva was usually associated with a particular individual, we found it simpler to inform the donor and request an adjustment to their collection method, such as providing a sample at a different time of the day or rinsing the mouth with water briefly before collection. This behavioral change improved the quality of the sample and substantially decreased the inconclusive rate. We also discovered that pooling saliva prior to testing can be beneficial in cases where a particular specimen is problematic due to low pH, color, or viscosity, or has substances that interfere with nucleic acid amplification. Pooling saliva for large-scale surveillance programs has proven to be a highly cost-effective strategy [12, 17, 38, 55, 56] and used successfully in schools and universities to routinely identify asymptomatic individuals in pool sizes comprised of 24 samples [17]. These studies include an RNA purification step and RT-qPCR for detection. We demonstrate here the successful detection of a single positive specimen in a pool size of 16 samples using the extraction-free RT-LAMP method.

To further determine the efficiency of the color readout upon visual inspection, the color output of 3654 samples was scored by 3 different operators without knowledge of the corresponding fluorescence readout. The high concordance between the results obtained by the different operators and their overall agreement with the real-time fluorescent readouts highlights the versatility and robustness of the RT-LAMP method. The visual readout option does not require sophisticated equipment or highly skilled personnel and is well suited to a low-resource or field setting. Use of a colorimetric signal also enables absorbance-based measurements of color (Color SARS-CoV-2 RT-LAMP Diagnostic Assay) and simple at home testing (Lucira™ COVID-19 Test Kit, Lucira Health), expanding the utility of RT-LAMP to be compatible with laboratories or settings without fluorescence instruments.

For reporting purposes, we used the fluorescent readout with a cutoff Tt < 26 minutes to determine positivity. We noted that in the case of positive samples, if the initial Tt is less than 11 minutes, all triplicate samples tested positive with a similar Tt value. More variance was observed in replicate testing at higher Tt values (data not shown). On a few occasions, either at the onset or end of infection, an inconclusive determination was made after the initial test scored positive but not all replicates tested positive. This likely reflected a low viral load present. We also noted more variance in the testing outcome late in infection with the same individual testing positive after receiving one or more consecutive negative test results, likely due to low levels of virus present in the saliva. This is consistent with the findings in our validation study showing that a viral load higher than 40 copies/μL of saliva can be detected confidently but samples with a lower viral titer would less likely be detected. In the clinical validation studies, overall sensitivity of 97% and specificity of 100% were achieved.

The high performance of the extraction-free RT-LAMP method was also demonstrated when compared with RT-qPCR using purified RNA as input material. Only samples with very late Cq values (greater than 34) in RT-qPCR, failed to amplify in extraction-free RT-LAMP. High Cq values (>34) in RT-qPCR have been shown to correlate with low viral loads (<100-1,000 copies/μL) and these individuals are rarely infectious or not infectious [41].

In an analysis of almost 100,000 individuals in the United States, more males tested positive for SARS-CoV-2 than females [57]. Viral loads have also been reported to be ∼ 10 times higher in males compared to females, as well as a slower viral clearance in males [30]. In our study, involving more than 30,000 saliva samples from 406 males, 341females and 8 gender not indicated, we did not observe a significant difference in positivity rates between genders, likely due to the small number of cases identified (47 males and 35 females). Interestingly, we found a higher viral load in samples from males, however, clearance rates did not differ between males and females. At 10 days after the initial positive test, we found 13% of positive individuals still testing positive with most of them testing negative for viral RNA by day 14. This may be a general trend for saliva-based COVID diagnostics since in a mass screening program in Slovenia, the viral load in saliva peaked within the first week of infection with most individuals testing negative within 2 weeks after the first positive RT-LAMP test [38].

During our surveillance, the peaks in new cases largely following a holiday period as has been reported in other testing programs [17], and generally mirrored the epidemiological data from Massachusetts and nationwide. The rapid testing time allowed the identification and isolation of positive individuals within hours of sample collection and resulted in low workplace transmission. The simple saliva workflow described requires minimal sample manipulation and is applicable to a variety of settings, budgets, scales of testing, and range of available infrastructure. It enables widespread and frequent diagnostic testing which is key to preventing the spread of SARS-CoV-2 infection in the population.

## Data Availability

All data produced in the present work are contained in the manuscript.

## Acknowledgments

This work was supported by New England Biolabs. We thank Amit Sinha and Sofia Roitman for participation in the blinded reading of colorimetric data, and Nicole Nichols for helpful discussions, Lori Tonello for organizing the many volunteers involved in saliva kit assembly, Tasha José for the diagrammatic representation of the workflow, Dr. Gyorgy Abel for medical oversight, Lea Antonopoulos for reagents, and Tom Evans for guidance and feedback on the manuscript.

## Notes

### Competing Interest Statement

New England Biolabs (www.neb.com) has funded this study. All authors (ZL,JLB, BC, CVC, WEJ, KK, BWL, JB, RAM, CBP, GR, RJR, NAT, YZ and CKSC) are employees and shareholders of New England Biolabs, manufacturer of LAMP and PCR reagents described in the manuscript

### Funding Statement

This study was funded by New England Biolabs.

### Author Declarations

Institutional Review Board (IRB)of WCG/New England Biolabs gave ethical approval for this work.

