## Supplementary File for "Development and Implementation of a Simple and Rapid Extraction-Free Saliva SARS-CoV-2 RT-LAMP Workflow for Workplace Surveillance"

### Supplementary Data

Figure S1. Heat treatment on endogenous actin detection in saliva. Contrived samples spiked with 2-10,000 viral copies/μL, were processed in triplicate using either 75ºC for 15 min, 85ºC for 10 min, 95ºC for 5 min, or no heat. The scanned image of the post-amplification actin LAMP plate with overlaid Tt values (upper panel) and the plot of actin Tt values (lower panel) are shown.

Figure S2. Suitability of the extraction-free RT-LAMP method for pooled saliva testing. Positive saliva was combined with equal volumes of 1 to 15 randomly selected negative saliva samples from different individuals, corresponding to dilution series of 1:2 through 1:16 of the original viral titer, and tested in COVID and actin LAMP reactions. All samples were tested in triplicate. LAMP Tt values are plotted.

**Figure S3. Sensitivity and specificity test.** Previously identified SARS-CoV-2 positive (n=30) and negative (n=30) samples were tested blindly in COVID and actin LAMP reactions. The scanned images of the post-amplification COVID and actin LAMP plates are shown with overlaid Tt values. Positive samples were revealed after the experiment and marked with red circles. The missed positive sample was marked with a red square. Negative and positive controls are marked with a blue rectangle. Experiments were performed 3 times with 2 different operators. The result from one experiment is shown.

**Figure S4. RT-qPCR standard curve using Twist RNA.** Different copy numbers of Twist RNA (5, 10, 100, 1000, and 10000) were used to generate a standard curve using the Luna® SARS-CoV-2 RT-qPCR Multiplex Assay Kit. Amplification and standard curves from both N1 and N2 targets are shown.

**Figure S5. Laboratory Information Management System: LIMS. (A)** User Interface: (1) Patient Log In, (2) Sample Submission and (3) Patient Result. **(B)** Operator Interface: (1) New Run, (2) Results Uploaded, (3) Sample Status, (4) Results Review and (5) Batch History.

**Table S1. Internal actin control detection of the 16 samples**

| | Purified Saliva RNA | | | | Saliva Lysate | |
|---|---|---|---|---|---|---|
| **Samples** | **RT-qPCR RNase P (Cq)** | | **Actin LAMP (Tt)** | | **Actin LAMP (Tt)** | |
| **1** | N/A | N/A | 8.9 | 9.3 | 9.1 | 9.1 |
| **2** | 43.1 | 43.2 | 10.4 | 10.3 | 11.1 | 11.2 |
| **3** | 30.4 | 30.2 | 7.2 | 7.2 | 8.8 | 8.8 |
| **4** | 33.9 | 39.5 | 9.2 | 9.3 | 10.6 | 10.6 |
| **5** | 41.1 | 39.0 | 8.3 | 8.3 | 11.1 | 11.4 |
| **6** | 35.1 | 34.8 | 8.5 | 8.7 | 12.1 | 12.0 |
| **7** | 35.4 | 35.9 | 9.4 | 9.4 | 10.2 | 10.3 |
| **8** | 33.7 | 33.2 | 8.3 | 8.3 | 11.8 | 11.9 |
| **9** | 40.1 | N/A | 10.8 | 11.5 | 10.7 | 10.7 |
| **10** | N/A | 38.0 | 11.0 | 11.8 | 10.6 | 10.4 |
| **11** | 39.9 | 37.3 | 9.1 | 9.0 | 9.4 | 9.9 |
| **12** | 33.2 | 33.7 | 9.7 | 9.6 | 9.6 | 9.7 |
| **13** | 33.7 | 33.7 | 8.4 | 8.5 | 10.3 | 10.2 |
| **14** | 37.7 | 38.9 | 10.8 | 11.1 | 10.6 | 10.0 |
| **15** | 32.8 | 32.9 | 8.5 | 8.5 | 10.0 | 10.0 |
| **16** | 38.8 | 38.8 | 9.8 | 10.2 | 10.5 | 10.5 |

No amplification is denoted N/A.

### Figure S1

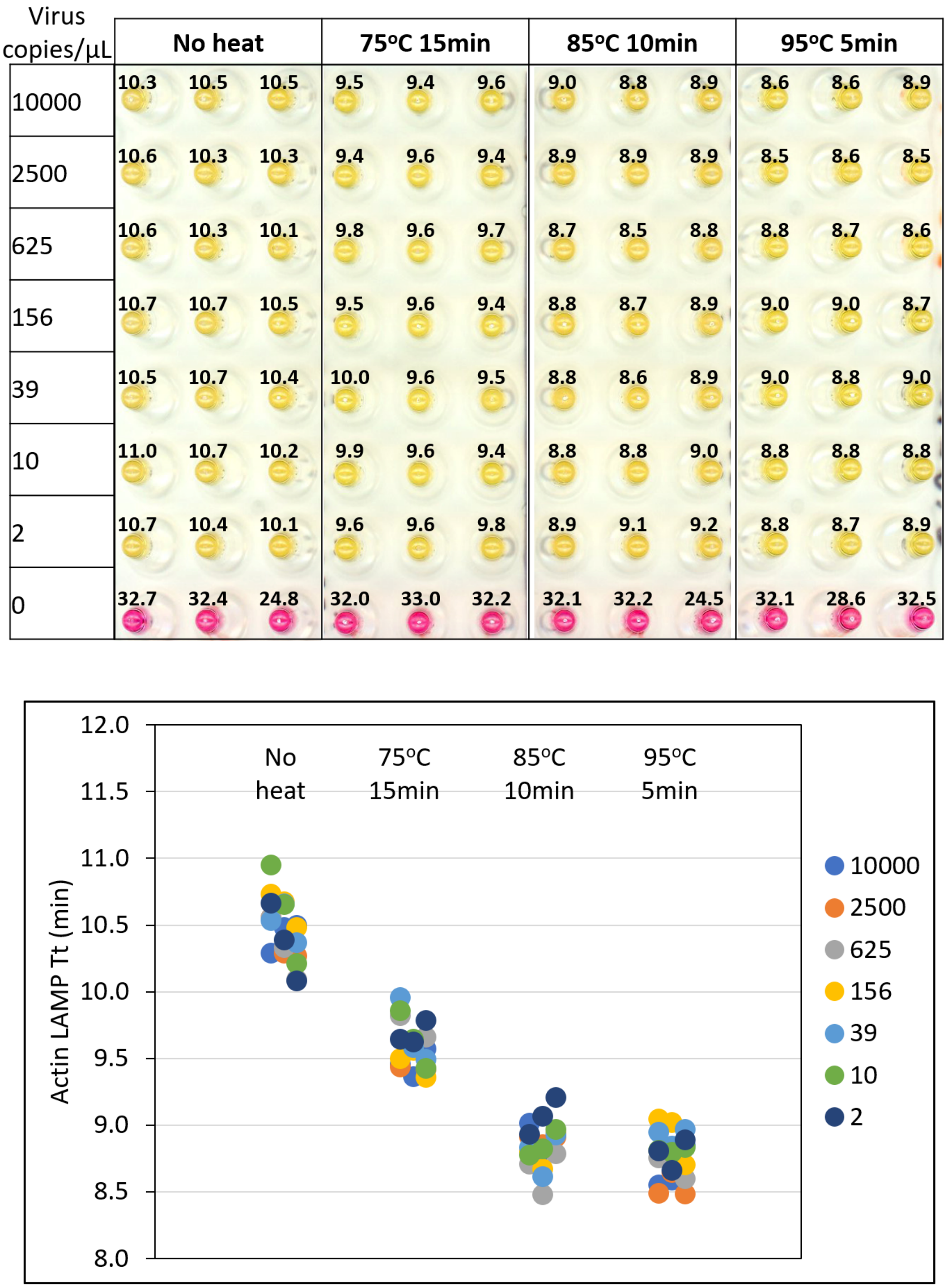
Virus copies/µL
No heat
75ºC 15min
85ºC 10min
95ºC 5min
10000 10.3 10.5 10.5 9.5 9.4 9.6 9.0 8.8 8.9 8.6 8.6 8.9
2500 10.6 10.3 10.3 9.4 9.6 9.4 8.9 8.9 8.9 8.5 8.6 8.5
625 10.6 10.3 10.1 9.8 9.6 9.7 8.7 8.5 8.8 8.8 8.7 8.6
156 10.7 10.7 10.5 9.5 9.6 9.4 8.8 8.7 8.9 9.0 9.0 8.7
39 10.5 10.7 10.4 10.0 9.6 9.5 8.8 8.6 8.9 9.0 8.8 9.0
10 11.0 10.7 10.2 9.9 9.6 9.4 8.8 8.8 9.0 8.8 8.8 8.8
2 10.7 10.4 10.1 9.6 9.6 9.8 8.9 9.1 9.2 8.8 8.7 8.9
0 32.7 32.4 24.8 32.0 33.0 32.2 32.1 32.2 24.5 32.1 28.6 32.5
Actin LAMP Tt (min)
12.0
11.5
11.0
10.5
10.0
9.5
9.0
8.5
8.0
No heat
75ºC 15min
85ºC 10min
95ºC 5min
10000
2500
625
156
39
10
2

### Figure S2

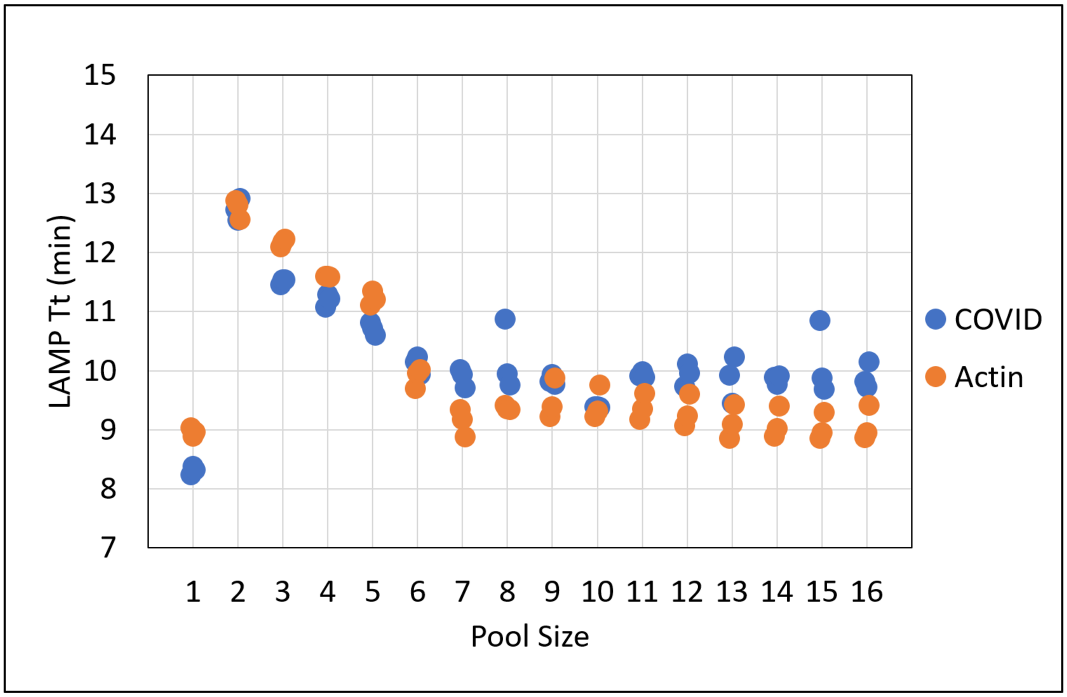


### Figure S3

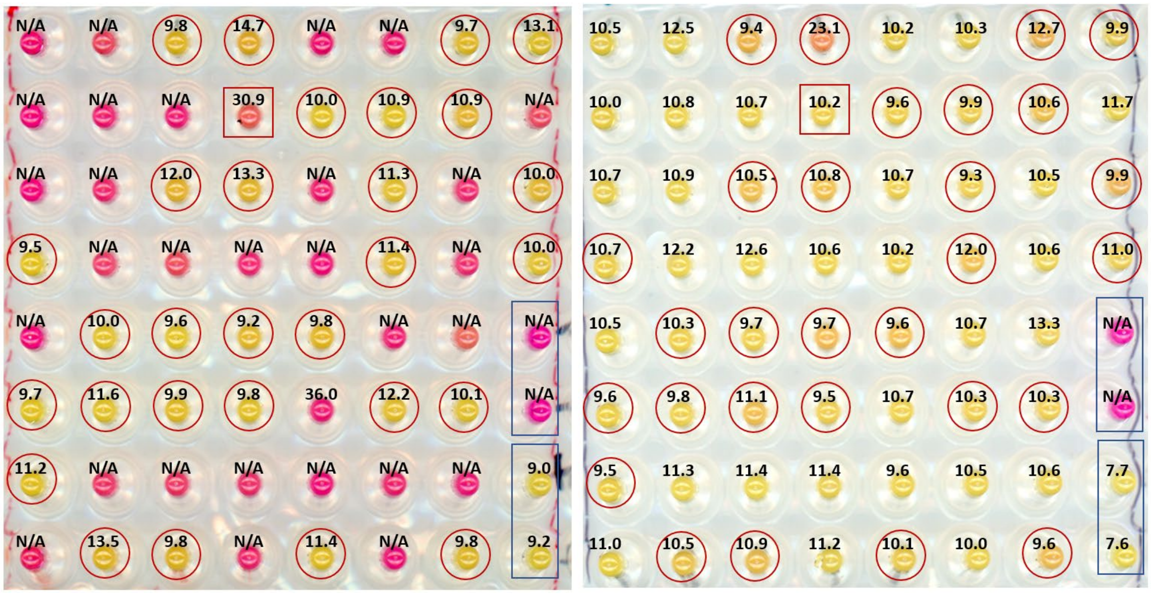

### Figure S4

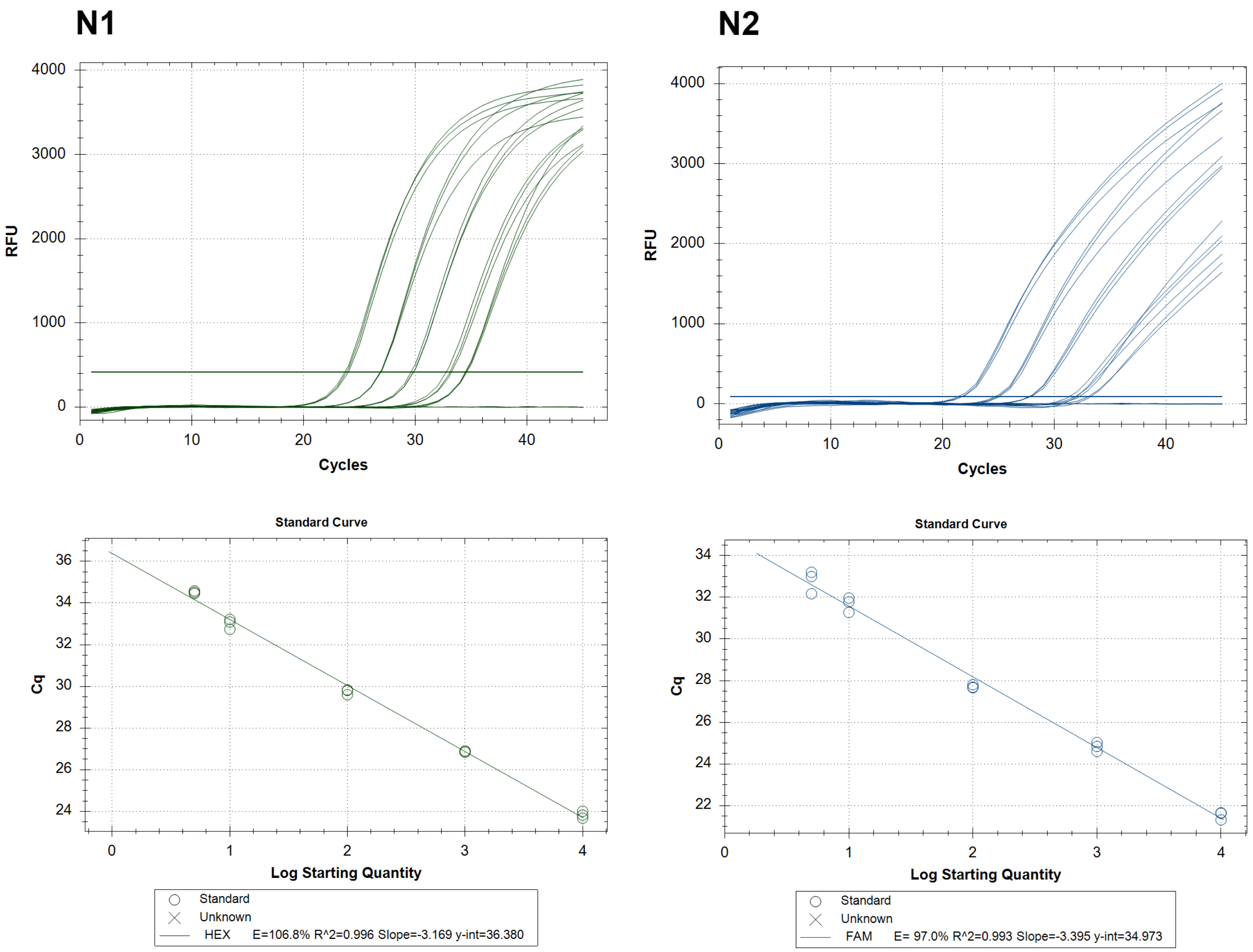

N1
N2
RFU
Cycles
Standard Curve
Cq
Log Starting Quantity
Standard
Unknown
HEX E=106.8% R^2=0.996 Slope=-3.169 y-int=36.380
FAM E= 97.0% R^2=0.993 Slope=-3.395 y-int=34.973

### Figure S5

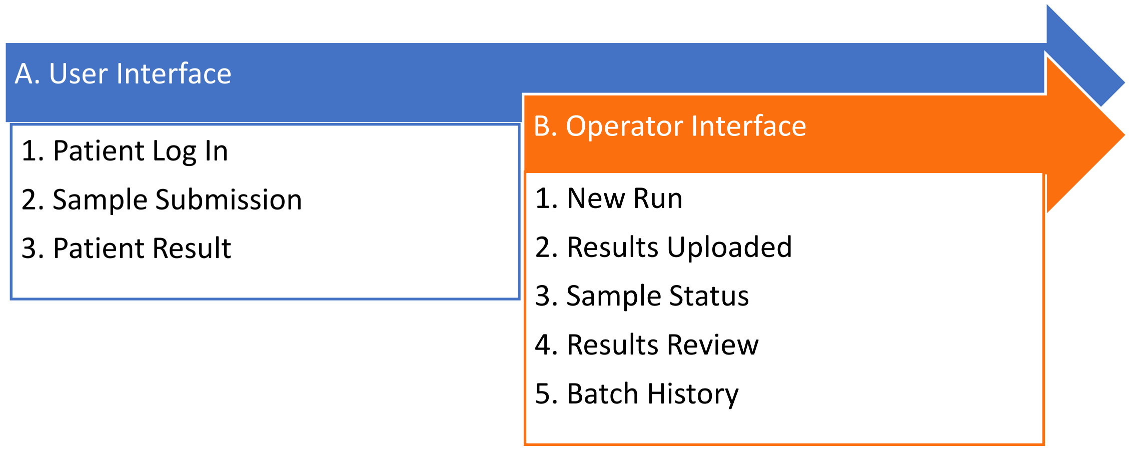


#### A. User Interface

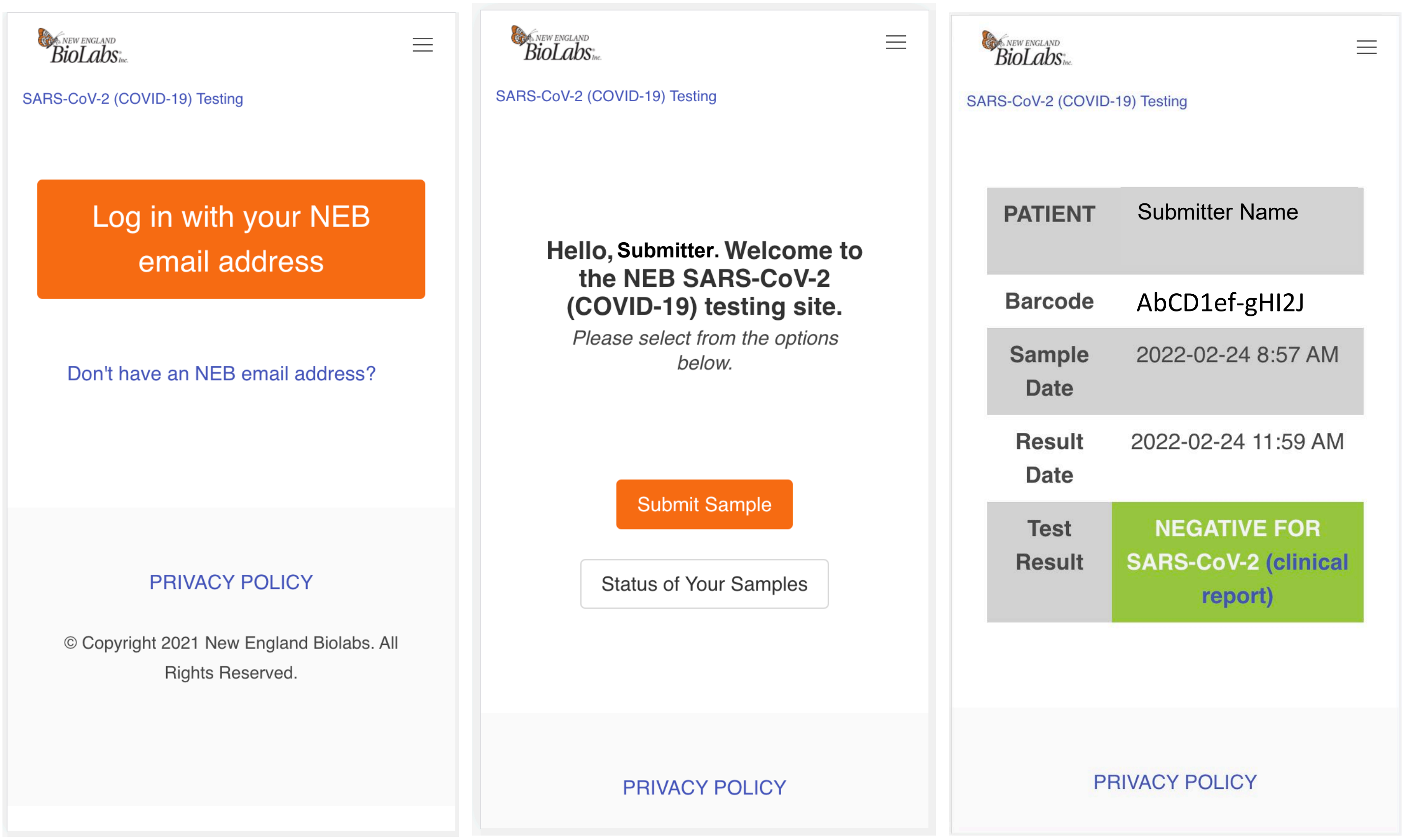

#### B. Operator Interface

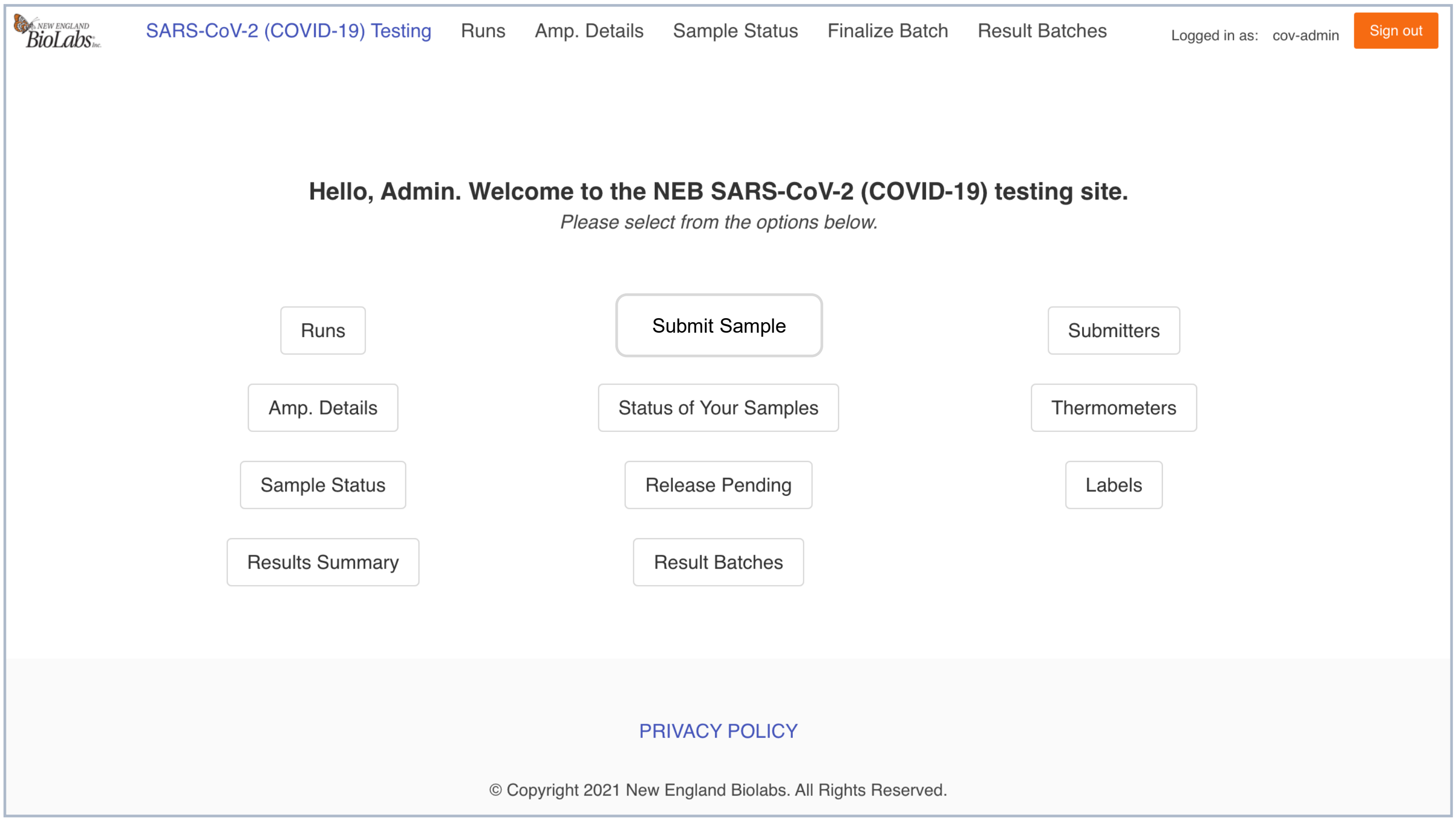


#### B1. New Run

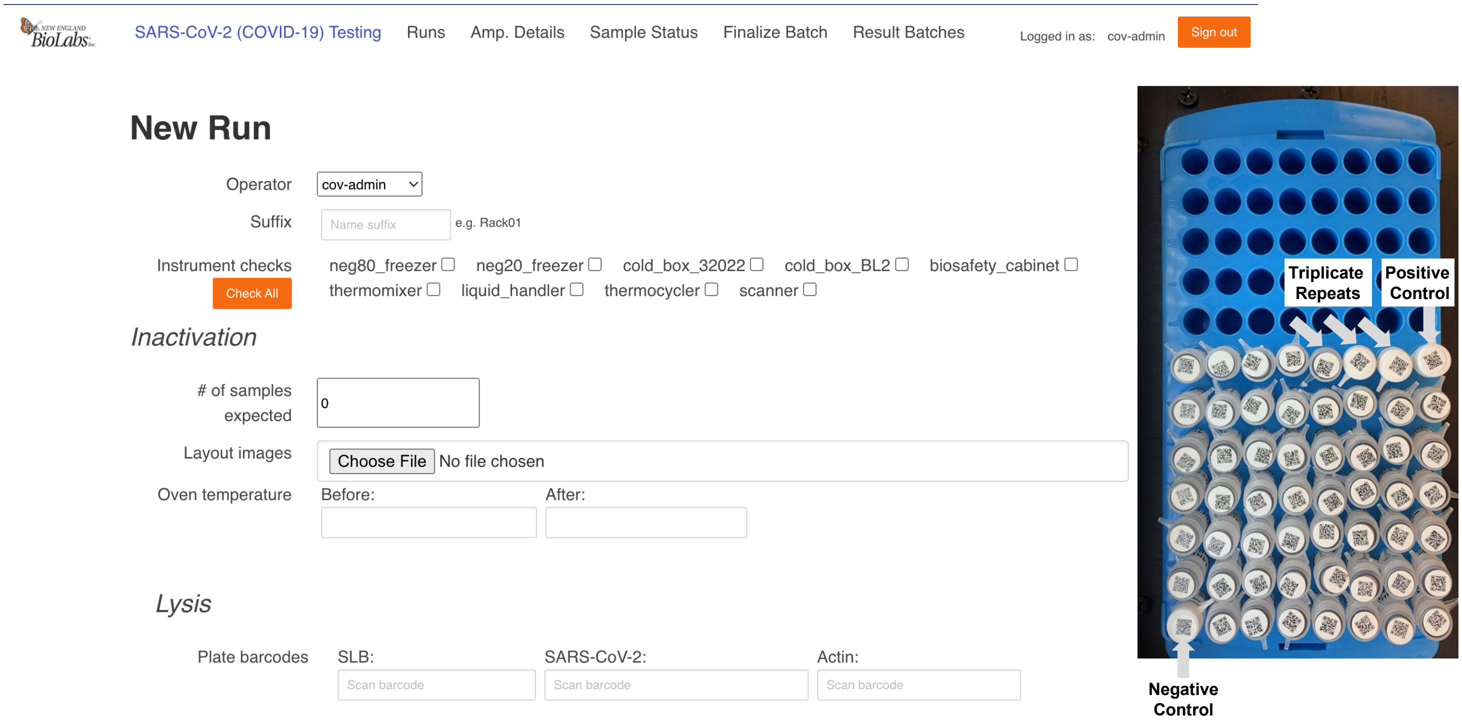

#### B2. Results Upload

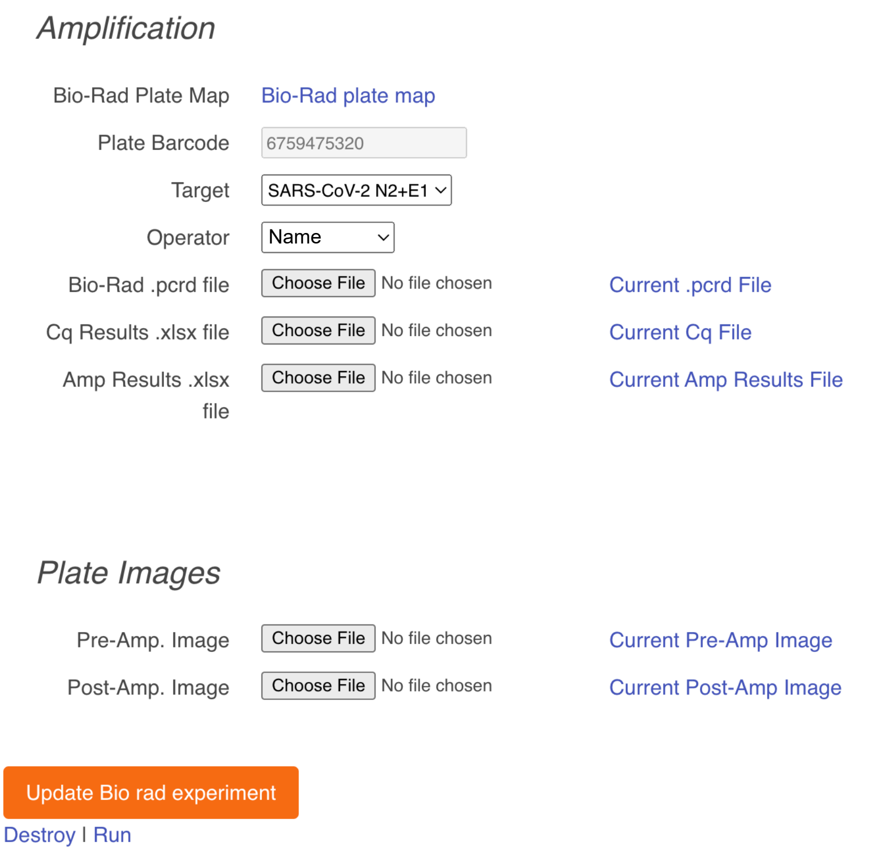


#### B3. Sample Status

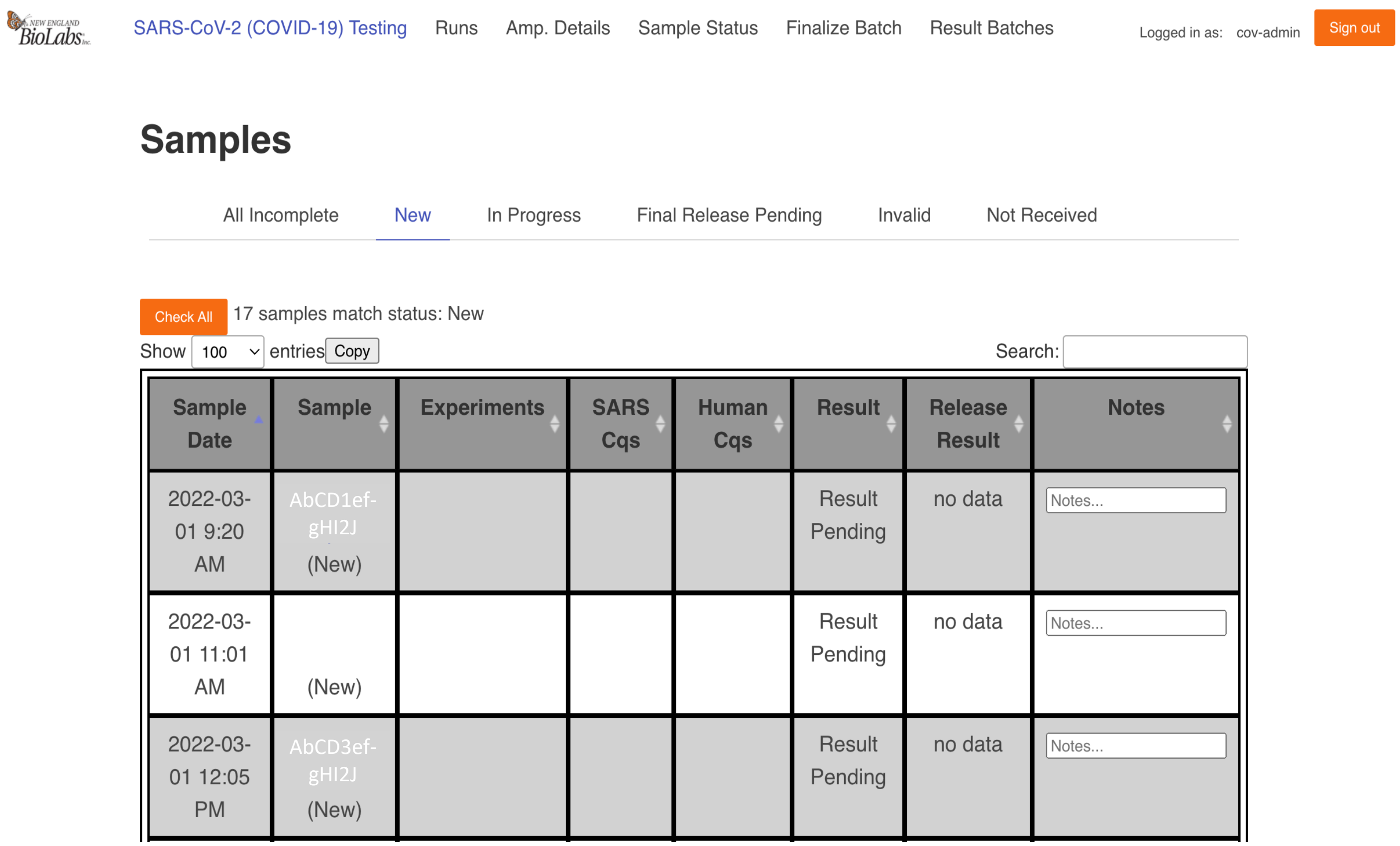

#### B4. Result Review

SARS-CoV-2 (COVID-19) Testing | Runs | Amp. Details | Sample Status | Finalize Batch | Result Batches

Logged in as: cov-admin | Sign out

### Release Pending Results

Temperature log: Choose File | Temperature logs (107).xlsx

Uncheck All

Show 1,000 entries | Copy

Search:

| Sample Registration Date | Sample | SARS-CoV-2 Cq Values | Human Cq Values | Result | Confirm | Notes |
|---|---|---|---|---|---|---|
| 2022-03-02 1:11 PM | AbCD1ef-gHI2J | N/A | 27.3 | not_detected | ☑ | |
| 2022-03-02 3:56 PM | | N/A | 25.3 | not_detected | ☑ | |
| 2022-03-02 3:58 PM | AbCD3ef-gHI2J | N/A | 25.6 | not_detected | ☑ | |
| 2022-03-02 4:03 PM | | N/A | 28.0 | not_detected | ☑ | |

#### B5. Batch History

SARS-CoV-2 (COVID-19) Testing | Runs | Amp. Details | Sample Status | Finalize Batch | Result Batches

Logged in as: cov-admin | Sign out

### SARS-CoV-2 Testing Results

Show 10 entries | Copy

Search:

| Batch Approval Date | Result Summary |
|---|---|
| 2022-03-03 | SARS_CoV-2 detected in 1/442 samples (0 inconclusive, 3 not identified) |
| 2022-03-01 | SARS_CoV-2 detected in 1/418 samples (0 inconclusive, 1 not identified) |
| 2022-02-28 | SARS_CoV-2 detected in 0/434 samples (0 inconclusive, 3 not identified) |
| 2022-02-24 | SARS_CoV-2 detected in 1/415 samples (0 inconclusive, 4 not identified) |
| 2022-02-22 | SARS_CoV-2 detected in 0/368 samples (0 inconclusive, 1 not identified) |
| 2022-02-18 | SARS_CoV-2 detected in 0/68 samples (0 inconclusive, 0 not identified) |
| 2022-02-17 | SARS_CoV-2 detected in 1/423 samples (0 inconclusive, 5 not identified) |
| 2022-02-15 | SARS_CoV-2 detected in 2/407 samples (0 inconclusive, 2 not identified) |
| 2022-02-14 | SARS_CoV-2 detected in 1/411 samples (0 inconclusive, 4 not identified) |
